# Prognostic impact of late left ventricular reverse remodeling in heart failure with reduced ejection fraction

**DOI:** 10.1101/2025.10.10.25337780

**Authors:** Hayao Ikesugi, Shinya Fujiki, Satomi Kawai, Sho Hirayama, Soma Sato, Kazuyo Tanaka, Yuka Sekiya, Hiroki Tsuchiya, Takayuki Kumaki, Ryohei Sakai, Hiromi Kayamori, Tsugumi Takayama, Takeshi Kashimura, Takayuki Inomata

**Affiliations:** Department of Cardiovascular Medicine, Niigata University Graduate School of Medicine, Dentistry and Health Sciences, Niigata, Japan

**Keywords:** heart failure, ejection fraction, left ventricular reverse remodeling, outcomes, guideline-directed medical therapy

## Abstract

Left ventricular reverse remodeling (LVRR) is associated with favorable outcomes in heart failure with reduced ejection fraction (HFrEF), but the prognostic significance of LVRR first documented beyond 24 months has not been fully characterized in patients with diverse etiologies. We retrospectively studied 213 patients with HFrEF (baseline left ventricular ejection fraction [LVEF] ≤40%) who underwent echocardiography at baseline and in two prespecified follow-up windows: 6 to <24 months (Follow-up 1) and 24–78 months (Follow-up 2). LVRR was defined as an absolute increase in LVEF of ≥10 percentage points and a follow-up LVEF >40%. Patients were classified as Early LVRR (first meeting the criteria at Follow-up 1; n = 84), Late LVRR (first meeting the criteria at Follow-up 2; n = 25), or No-LVRR (not meeting the criteria at either follow-up; n = 104). The primary endpoint was a composite of all-cause mortality and heart failure hospitalization, assessed from Follow-up 2. Over a median follow-up of 33.8 months, 52 patients died and 32 were hospitalized for heart failure. In multivariable Cox regression adjusted for age, sex, and ischemic heart disease, both Early and Late LVRR were associated with a lower risk of the primary endpoint than No-LVRR (hazard ratio [HR] 0.299, 95% confidence interval [CI] 0.164–0.544, and HR 0.293, 95% CI 0.115–0.744, respectively). These associations remained significant in alternative models adjusted for diabetes mellitus, estimated glomerular filtration rate, or heart failure duration. Kaplan–Meier analyses showed better event-free survival for the composite endpoint and higher overall survival in both LVRR groups than in the No-LVRR group, whereas significantly better HF hospitalization-free survival was observed only in the Early LVRR group. The mean Simple-GDMT Score, reflecting guideline-directed medical therapy (GDMT) intensity, increased from baseline to Follow-up 1 in all three groups, but a further significant increase from Follow-up 1 to Follow-up 2 was observed only in the Late LVRR group. In multivariable linear regression, greater increases in GDMT intensity were associated with higher LVEF at Follow-up 2 (β 1.265, 95% CI 0.237–2.293). Late LVRR was associated with favorable clinical outcomes, and greater longitudinal GDMT intensification was associated with higher LVEF at Follow-up 2, although causal relationships cannot be established from this observational study.

## Introduction

Heart failure (HF) is a growing global health challenge, with rising prevalence and mortality [1, 2]. It represents the terminal stage of many cardiovascular diseases and is commonly classified by left ventricular ejection fraction (LVEF). Heart failure with reduced ejection fraction (HFrEF), typically defined as LVEF ≤40%, carries particularly high risks of death and hospitalization [3]. Guideline-directed medical therapy (GDMT), including ACE inhibitors (ACEis), angiotensin receptor blockers (ARBs), angiotensin receptor–neprilysin inhibitors (ARNi), β-blockers (BBs), mineralocorticoid receptor antagonists (MRAs), and sodium–glucose cotransporter 2 inhibitors (SGLT2is) has improved outcomes in patients with HFrEF. Selected non-pharmacological interventions, such as cardiac resynchronization therapy and coronary revascularization, also improve outcomes in appropriately selected patients [4]. In addition to reducing mortality and hospitalization, effective treatment may promote structural and functional improvement of the left ventricle. Left ventricular reverse remodeling (LVRR), characterized by improvement in left ventricular systolic function and reduction in chamber size, is therefore considered an important marker of treatment response and is associated with a more favorable prognosis [5].

LVRR is generally observed within the first 24 months of treatment, and early achievement of LVRR is associated with a favorable prognosis [6]. Therefore, most previous studies have focused on the clinical significance of LVRR occurring within the first 24 months [7–9]. However, some patients may achieve LVRR beyond this timeframe [6]. LVRR occurring beyond 24 months may intuitively be regarded as a favorable sign of continued myocardial recovery. Indeed, such delayed LVRR has been associated with favorable outcomes in patients with idiopathic dilated cardiomyopathy [10]. However, its prognostic significance in patients with HFrEF of diverse etiologies has not been fully elucidated. Clarifying the clinical characteristics and prognostic significance of delayed LVRR may be particularly relevant in real-world practice, where optimization of treatment may be gradual and the timing of recovery may vary substantially among patients. Therefore, this study aimed to investigate the clinical features and prognostic relevance of late-onset LVRR (≥24 months) compared with early-onset LVRR (<24 months) and no LVRR.

## Materials and methods

### Study design and patient population

This single-center, retrospective observational study was conducted at Niigata University Medical and Dental Hospital. We included patients with HFrEF (LVEF ≤40%) who underwent baseline transthoracic echocardiography (TTE) between January 2011 and December 2022. Baseline was defined as the first clinically available TTE performed at our institution. All patients underwent two subsequent TTE examinations: Follow-up 1 (6 to <24 months) and Follow-up 2 (24–78 months). When multiple TTEs were available within a prespecified follow-up period, the earliest examination was selected.

For the purposes of this study, clinically manifest HF was operationally defined as a history of HF hospitalization or ongoing diuretic therapy (including loop diuretics, thiazide diuretics, or tolvaptan) at the time of the baseline TTE. Patients who met neither criterion were excluded [4]. At baseline, clinical information, including medical history, comorbidities, medications, and laboratory data, was collected from electronic medical records. HF duration was defined as the interval from the earliest documented diagnosis of HF in the medical records to the baseline TTE. At Follow-up 1 and Follow-up 2, echocardiographic data and data on pharmacological and non-pharmacological therapies were collected.

### Echocardiographic assessment and LVRR definition

All TTEs were performed under stable clinical conditions by certified sonographers in accordance with current echocardiographic guidelines [11, 12]. Echocardiographic assessments included LVEF, left ventricular dimensions, and the severity of valvular heart disease. Valvular heart disease was defined as moderate or greater aortic stenosis, aortic regurgitation, mitral regurgitation, or mitral stenosis according to standard echocardiographic guideline criteria [13].

LVRR was defined as an absolute increase in LVEF of ≥10 percentage points from baseline, with a follow-up LVEF >40% [14]. LVRR was classified according to the time at which it was first documented. Early LVRR was defined as meeting the criteria at Follow-up 1 (6 to <24 months), whereas Late LVRR was defined as meeting the criteria for the first time at Follow-up 2 (24–78 months). Patients who did not meet the LVRR criteria at either follow-up were categorized as No-LVRR.

### Pharmacological and non-pharmacological therapy assessment

Pharmacological and non-pharmacological therapies were assessed to characterize longitudinal treatment patterns in relation to the timing of LVRR. The prescription regimen documented at the clinical encounter closest to each echocardiographic assessment was used to calculate the Simple-GDMT Score **(Table 1)**, based on the type and dosage of evidence-based therapies for HFrEF, including RAS inhibitors (ACEis, ARBs, or ARNi), BBs, MRAs, and SGLT2is [15]. Although dosage standards in Japan differ from those in Europe and the United States because of differences in body size and pharmacological tolerability, the Simple-GDMT Score ranges from 0 to 9, reflecting the degree of GDMT optimization. The ΔSimple-GDMT Score, defined as the difference between the Follow-up 2 and baseline scores, was used to characterize longitudinal changes in GDMT intensity. Loop diuretic use was evaluated separately by converting prescribed doses to furosemide-equivalent doses using the following formula: 40 mg/day furosemide = 60 mg/day azosemide = 8 mg/day torsemide [16].

**Table 1.** Components and scoring of the Simple-GDMT Score.

| Simple GDMT score |  |  |
| --- | --- | --- |
| RASi | None | 0 |
|  | <50% max daily dose ACEi/ARB | 1 |
|  | ≥50% max daily dose ACEi/ARB | 2 |
|  | ARNi (any dose) | 3 |
| Beta-blocker | None | 0 |
|  | <50% max daily dose | 1 |
|  | ≥50% max daily dose | 2 |
| MRA | None | 0 |
|  | Any dose | 2 |
| SGLT2i | None | 0 |
|  | Any dose | 2 |
| total |  | 0-9 |

The Simple-GDMT Score was calculated as follows: RASi were scored 0 if not initiated, 1 if <50% of target dose, and 2 if 50–100% of target dose; ARNi were scored 3 regardless of dose; beta-blockers were scored 0 if not initiated, 1 if <50% of target dose, and 2 if 50–100% of target dose; and MRAs and SGLT2 inhibitors were scored 0 if not initiated and 2 if initiated regardless of dose. The total score ranged from 0 to 9.
Abbreviations: ACEi, angiotensin-converting enzyme inhibitor; ARB, angiotensin II receptor blocker; ARNi, angiotensin receptor–neprilysin inhibitor; GDMT, guideline-directed medical therapy; MRA, mineralocorticoid receptor antagonist; RASi, renin–angiotensin system inhibitor; SGLT2i, sodium–glucose cotransporter 2 inhibitor.

Similarly, we recorded the rates of the following procedures: pacemaker implantation, implantable cardioverter-defibrillator (ICD) implantation, cardiac resynchronization therapy with defibrillator (CRT-D), percutaneous coronary intervention (PCI), catheter ablation, coronary artery bypass grafting (CABG), and valve intervention.

### Outcomes

Clinical outcomes, including hospitalizations and deaths, were assessed from the date of the Follow-up 2 TTE examination, in a landmark analysis framework. Patients were followed for up to five years thereafter. The primary endpoint was a composite of all-cause mortality and heart failure hospitalization. Secondary endpoints included each component of the primary endpoint analyzed separately: all-cause mortality and heart failure hospitalization.

### Statistical analysis

Continuous variables were expressed as mean ± standard deviation or median [interquartile range], as appropriate, and categorical variables as number (percentage). Baseline characteristics among the three LVRR groups, including the Simple-GDMT Score (analyzed as a continuous variable), were compared using the Kruskal–Wallis test for continuous variables and the chi-square test for categorical variables. For post hoc pairwise comparisons, the Mann–Whitney U test and Fisher’s exact test were applied to continuous and categorical variables, respectively.

Longitudinal changes in continuous variables, including echocardiographic parameters, the Simple-GDMT Score, and loop diuretic doses, were assessed using the Friedman test, followed by the Wilcoxon signed-rank test for pairwise comparisons within each group. Longitudinal changes in categorical treatment variables, including pharmacological and non-pharmacological therapies, were assessed using Cochran’s Q test, followed by McNemar’s test for pairwise comparisons. To identify factors associated with LVEF at Follow-up 2, multivariable linear regression analyses were performed. Based on clinical relevance and previous studies of cardiac remodeling and LVEF, age, sex, HF duration, ischemic heart disease, baseline LVEF, estimated glomerular filtration rate (eGFR), and brain natriuretic peptide (BNP) were included a priori as covariates [17–19]. BNP was log-transformed because of its skewed distribution. The ΔSimple-GDMT Score was included as a variable of clinical interest to examine the association between changes in GDMT intensity and LVEF at Follow-up 2. Model 1 included the ΔSimple-GDMT Score, baseline LVEF, age, and sex. Model 2 additionally included ischemic heart disease, log-transformed BNP, eGFR, and HF duration. HF duration was modeled using restricted cubic splines with three knots.

Survival analyses were performed using Kaplan–Meier estimates, and group differences were assessed with the log-rank test. Patients were censored at the time of the last clinical contact if follow-up ended due to transfer of care, whereas patients who remained under follow-up without events were administratively censored at the end of the observation period. To examine the association between LVRR classification and clinical outcomes, multivariable Cox proportional hazards models were constructed with the LVRR group as the primary independent variable. Covariates were selected a priori based on clinical relevance and prior evidence. Model 1 was adjusted for age, sex, and ischemic heart disease; Model 2 for age, sex, and diabetes mellitus; Model 3 for age, sex, and renal function assessed by eGFR; and Model 4 for age, sex, and HF duration. Follow-up time was measured from the date of the Follow-up 2 TTE. For pairwise comparisons among the three LVRR groups and longitudinal pairwise comparisons across the three time points within each group, a Bonferroni-corrected significance threshold of p < 0.017 was applied. The same threshold was applied to pairwise log-rank tests. For other analyses, a two-sided p < 0.05 was considered statistically significant. Statistical analyses were performed using IBM SPSS Statistics version 25.0 (IBM Corp., Armonk, NY, USA) and R version 4.6.1 (R Foundation for Statistical Computing, Vienna, Austria).

### Ethics statement

This study was conducted in accordance with the Declaration of Helsinki and was approved by the institutional ethics committee of Niigata University (Approval No. 2023-0351). The requirement for informed consent was waived due to the retrospective observational design. This study was registered in the University Hospital Medical Information Network (UMIN) Clinical Trials Registry (Registration No. UMIN000056411) as an observational study.

## Results

### Participants and baseline characteristics

Among 1049 patients with HFrEF who underwent baseline TTE between January 2011 and December 2022, 421 were excluded because the patients had no history of hospitalization for HF or ongoing diuretic therapy, 300 because of missing TTE data at 6 to <24 months (death, n = 52; other loss, n = 248), and 115 because of missing TTE data at 24–78 months (death, n = 29; other loss, n = 86). Other losses were mainly due to transfer of care or lack of echocardiographic follow-up within the prespecified windows. Ultimately, 213 patients were included: 84 (39.4%) with Early LVRR, 25 (11.7%) with Late LVRR, and 104 (48.8%) with No-LVRR **(Figure 1)**.

**Figure 1:**
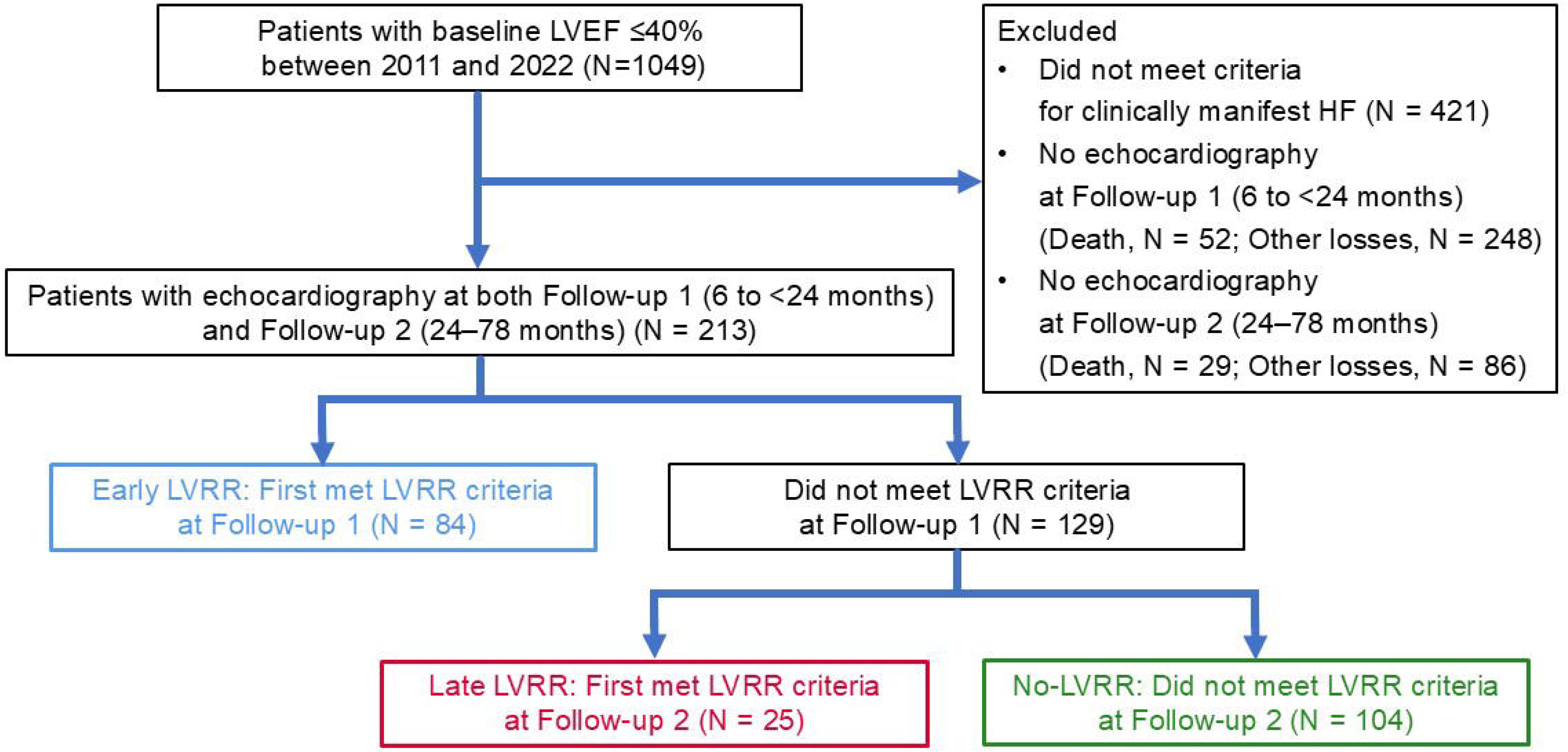
Flowchart of patient selection and LVRR classification Abbreviations: LVEF, left ventricular ejection fraction; LVRR, left ventricular reverse remodeling.

The mean age was 61.3 ± 13.6 years, 74.6% were male, and the mean LVEF was 28.5 ± 7.8%. Age, sex, and baseline LVEF did not differ significantly among the three groups. Ischemic etiology was least frequent in the Early LVRR group and most frequent in the No-LVRR group (p = 0.014), with the Late LVRR group showing intermediate characteristics. eGFR differed significantly among the three groups (overall p = 0.014), with higher values in the Late LVRR group than in the No-LVRR group (p = 0.009). HF duration before the baseline TTE differed between the groups, with the Late LVRR group having values between those observed in the Early LVRR and No-LVRR groups.

At baseline, the implementation rates of ICD, CRT-D, and PCI differed among the groups (overall p = 0.003, p = 0.001, and p = 0.001, respectively), with the Early LVRR group showing the lowest rates. The baseline Simple-GDMT Score differed among groups (overall p = 0.018), with a lower score in the Early LVRR group than in the No-LVRR group (3.1 ± 1.8 vs. 3.9 ± 1.7, p = 0.007). Similarly, the furosemide-equivalent loop diuretic dose was lower in the Early LVRR group than in the No-LVRR group (27.3 ± 15.3 vs. 39.2 ± 24.6, p < 0.001) **(Table 2)**.

**Table 2.**
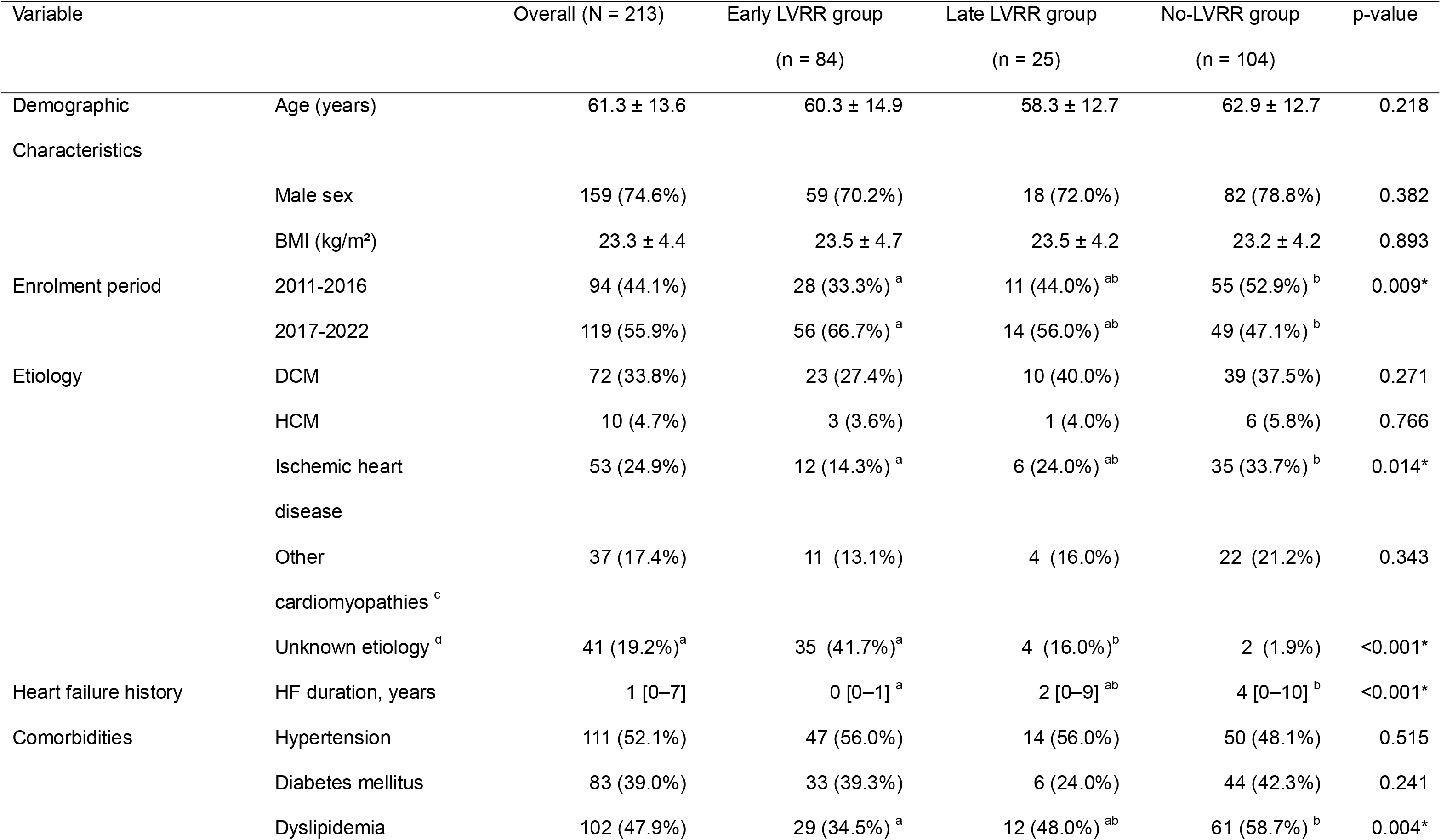
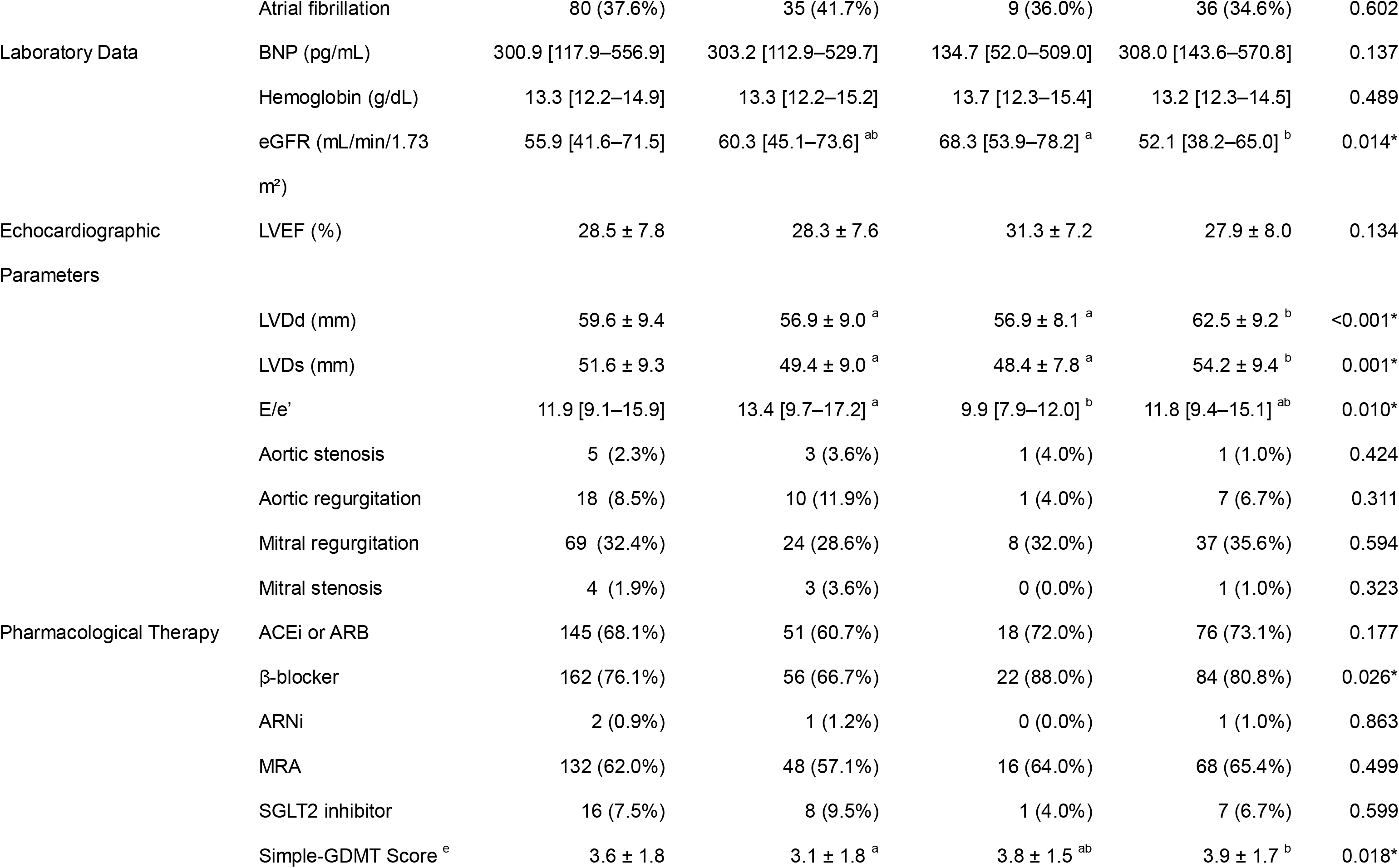
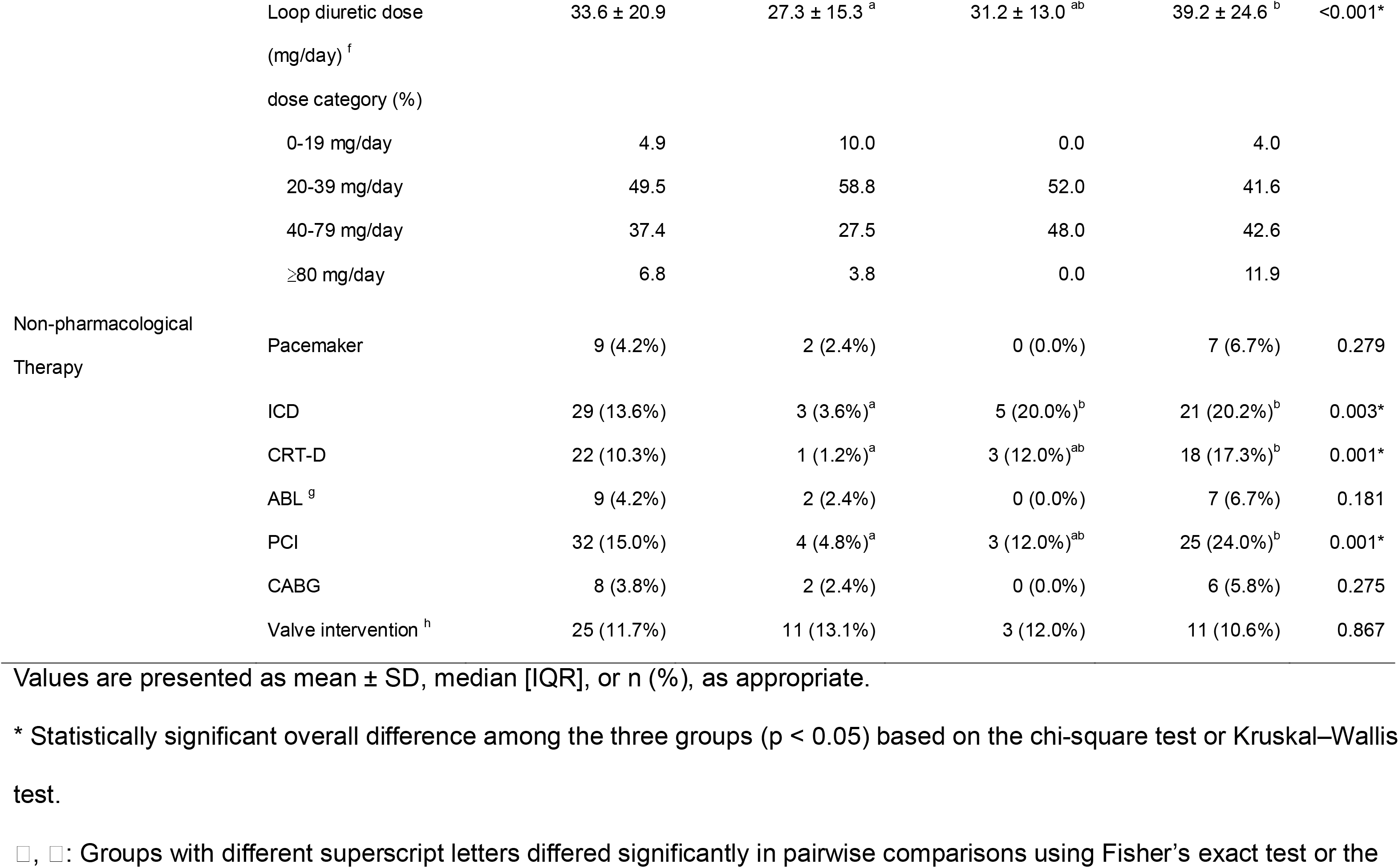
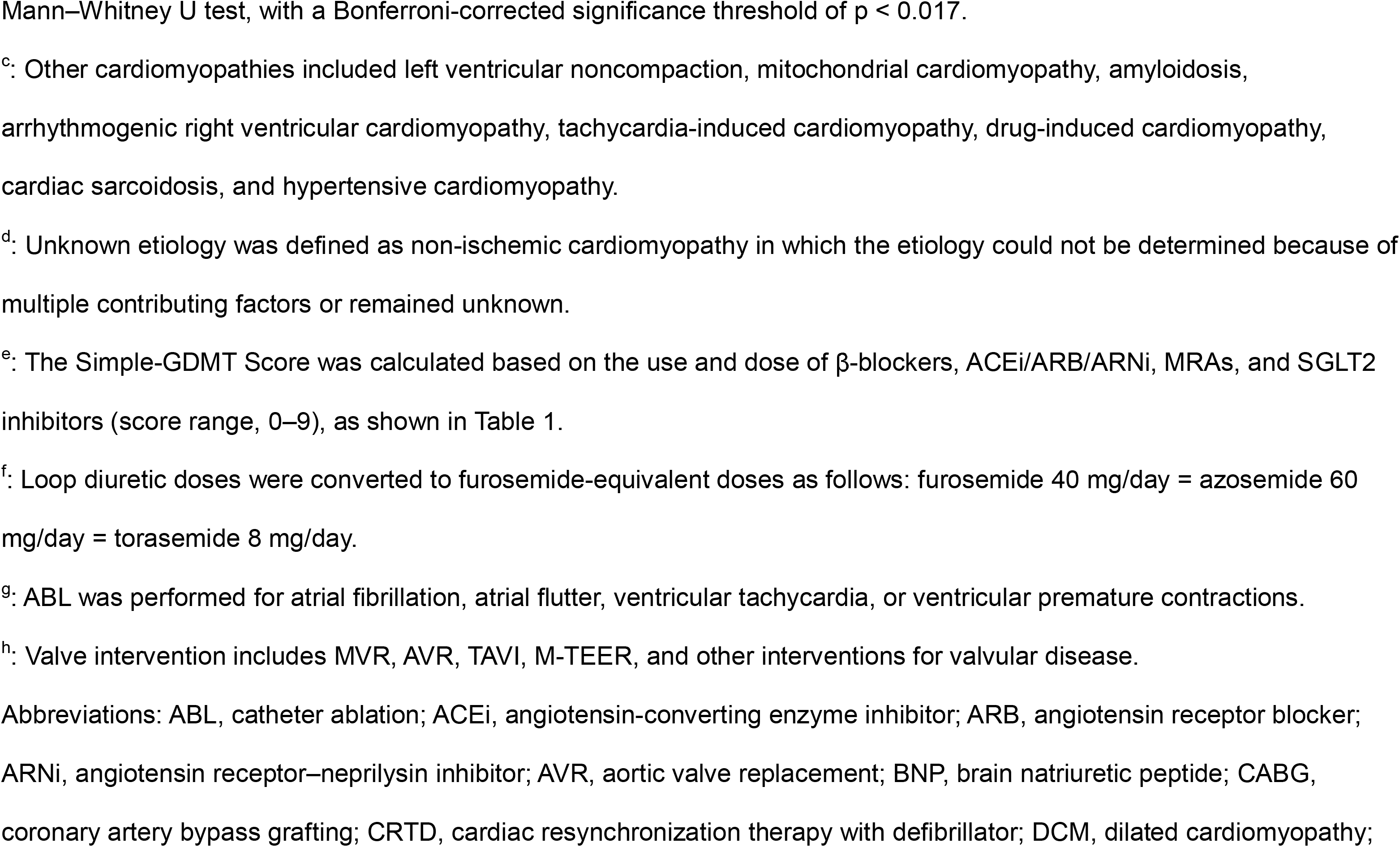
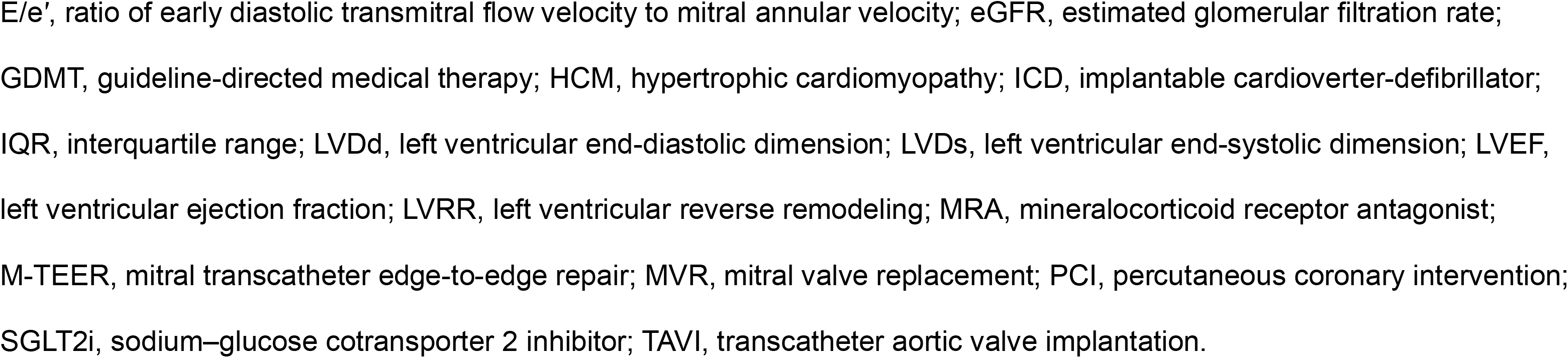
Baseline characteristics.

### Changes in TTE parameters over time

The median intervals from baseline to Follow-up 1 and Follow-up 2 were 365 and 914 days, respectively. The Early LVRR group exhibited marked reverse remodeling by Follow-up 1, with improvements in LVEF and reductions in LV dimensions that were maintained at Follow-up 2. In the Late LVRR group, remodeling was modest at Follow-up 1 but became substantial by Follow-up 2. In contrast, the No-LVRR group showed minimal changes over time (Supplementary Table S1 in Online Resource 1).

### Changes in pharmacological and non-pharmacological therapies

Pharmacological therapy was intensified over time in all three groups, as reflected by increases in the Simple-GDMT Score **(Figure 2)**. In the Early LVRR group, the Simple-GDMT Score increased from 3.1 ± 1.8 at baseline to 4.6 ± 1.9 at Follow-up 1 (p < 0.001), with no further significant increase by Follow-up 2 (4.7 ± 2.0, p = 0.224). In contrast, the Late LVRR group showed progressive increases in the score, from 3.8 ± 1.5 at baseline to 4.5 ± 1.4 at Follow-up 1 (p = 0.007) and 5.2 ± 1.9 at Follow-up 2 (p = 0.005 for Follow-up 1 vs. Follow-up 2). In the No-LVRR group, the score increased from 3.9 ± 1.7 at baseline to 4.3 ± 2.1 at Follow-up 1 (p = 0.010), but not further by Follow-up 2 (4.8 ± 2.4, p = 0.042). Only the Late LVRR group showed a significant increase in the Simple-GDMT Score across both follow-up intervals. Changes in implementation rates by drug class are shown in **Supplementary Figure S1 in Online Resource 1**. Loop diuretic prescription rates and furosemide-equivalent doses also changed during follow-up **(Supplementary Figure S2 in Online Resource 1)**. Regarding non-pharmacological therapies, the Early LVRR group showed significant increases in CRT-D implantation, PCI, catheter ablation, and valve intervention, with most changes occurring between baseline and Follow-up 1. In the Late LVRR group, CRT-D implantation increased over time (overall p = 0.011), although none of the pairwise comparisons reached statistical significance. In the No-LVRR group, several interventions, including CRT-D implantation, PCI, catheter ablation, and valve intervention, increased over the follow-up period **(Table 3)**.

**Figure 2:**
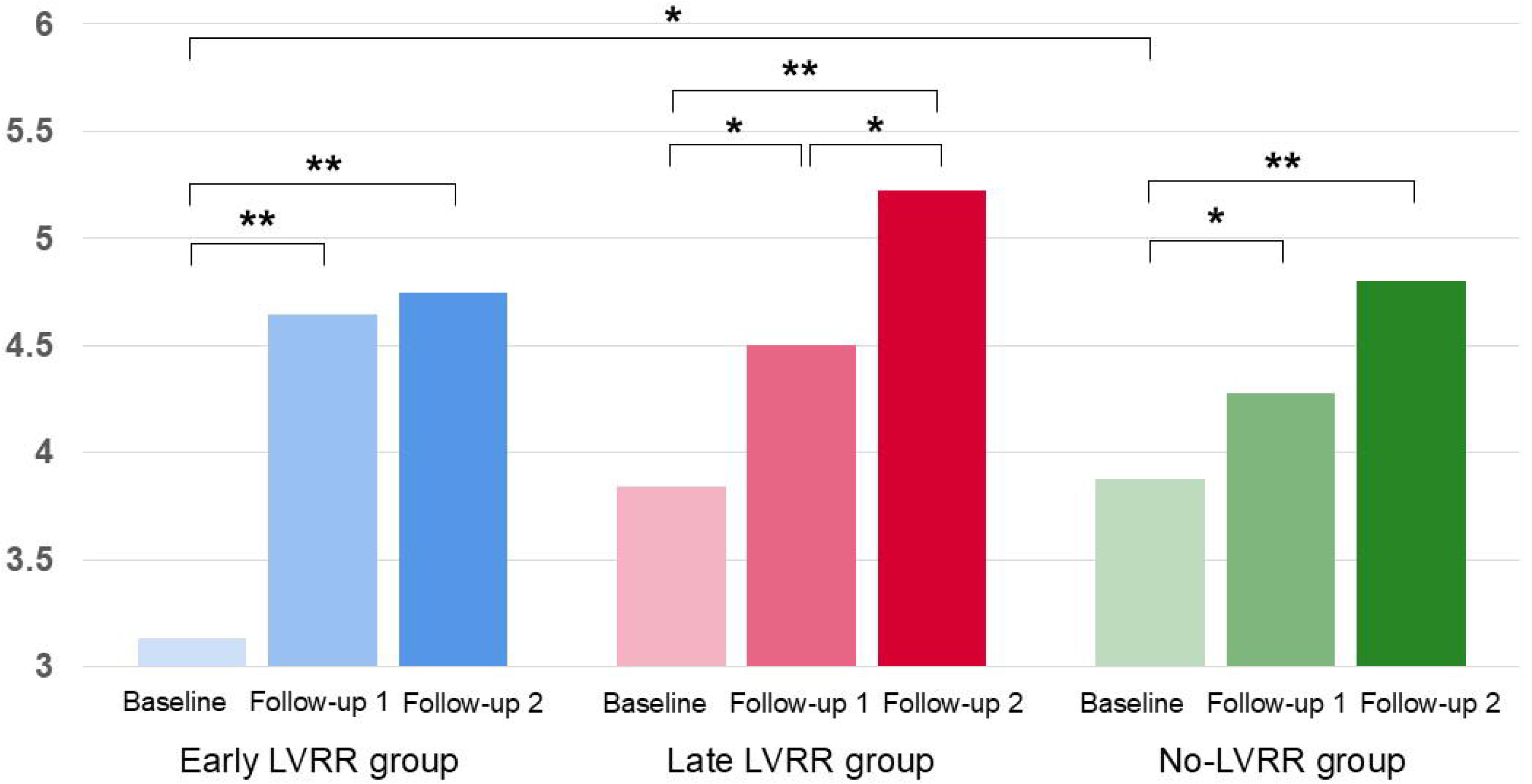
Trajectories of the Simple-GDMT Score in each LVRR group Blue bars represent the Early LVRR group, red bars the Late LVRR group, and green bars the No-LVRR group. Each bar represents the mean Simple-GDMT Score at baseline, Follow-up 1 (6 to <24 months), and Follow-up 2 (24–78 months). Brackets indicate pairwise comparisons either between time points within each LVRR group or between LVRR groups at the same time point. Within-group pairwise comparisons were performed using the Wilcoxon signed-rank test, and between-group pairwise comparisons were performed using the Mann–Whitney U test. *p < 0.017; **p < 0.0033. The Bonferroni-corrected significance threshold was p < 0.017. Abbreviations: GDMT, guideline-directed medical therapy; LVRR, left ventricular reverse remodeling.

**Table 3.** Longitudinal changes in non-pharmacological therapy from Baseline to Follow-up 2 according to LVRR status.

**Early LVRR group (n = 84)**
| Non-pharmacological Therapy | Baseline | Follow-up 1 | Follow-up 2 | p-value |
| --- | --- | --- | --- | --- |
| Pacemaker | 2 (2.4%) | 1 (1.2%) | 1 (1.2%) | 0.368 |
| ICD | 3 (3.6%) | 6 (7.1%) | 6 (7.1%) | 0.050 |
| CRT-D | 1 (1.2%) | 6 (7.1%) | 6 (7.1%) | 0.007* |
| PCI | 4 (4.8%)<sup>a</sup> | 11 (13.1%)<sup>b</sup> | 11 (13.1%)<sup>b</sup> | <0.001* |
| ABL <sup>c</sup> | 2 (2.4%)<sup>a</sup> | 9 (10.7%)<sup>b</sup> | 14 (16.7%)<sup>b</sup> | <0.001* |
| CABG | 2 (2.4%) | 2 (2.4%) | 2 (2.4%) | — |
| Valve intervention <sup>d</sup> | 11 (13.1%)<sup>a</sup> | 20 (23.8%)<sup>b</sup> | 20 (23.8%)<sup>b</sup> | <0.001* |

**Late LVRR group (n = 25)**
| Non-pharmacological Therapy | Baseline | Follow-up 1 | Follow-up 2 | p-value |
| --- | --- | --- | --- | --- |
| Pacemaker | 0 (0.0%) | 0 (0.0%) | 0 (0.0%) | — |
| ICD | 5 (20.0%) | 6 (24.0%) | 4 (16.0%) | 0.368 |
| CRT-D | 3 (12.0%) | 6 (24.0%) | 9 (36.0%) | 0.011* |
| PCI | 3 (12.0%) | 4 (16.0%) | 4 (16.0%) | 0.368 |
| ABL <sup>c</sup> | 0 (0.0%) | 2 (8.0%) | 2 (8.0%) | 0.135 |
| CABG | 0 (0.0%) | 2 (8.0%) | 2 (8.0%) | 0.135 |
| Valve intervention <sup>d</sup> | 3 (12.0%) | 5 (20.0%) | 5 (20.0%) | 0.135 |

**No LVRR group (n = 104)**
| Non-pharmacological Therapy | Baseline | Follow-up 1 | Follow-up 2 | p-value |
| --- | --- | --- | --- | --- |
| Pacemaker | 7 (6.7%) | 4 (3.8%) | 3 (2.9%) | 0.039* |
| ICD | 21 (20.2%) | 23 (22.1%) | 23 (22.1%) | 0.717 |
| CRT-D | 18 (17.3%)<sup>a</sup> | 36 (34.6%)<sup>b</sup> | 42 (40.4%)<sup>b</sup> | <0.001* |
| PCI | 25 (24.0%)<sup>a</sup> | 30 (28.8%)<sup>ab</sup> | 33 (31.7%)<sup>b</sup> | 0.002* |
| ABL <sup>c</sup> | 7 (6.7%)<sup>a</sup> | 12 (11.5%)<sup>ab</sup> | 17 (16.3%)<sup>b</sup> | <0.001* |
| CABG | 6 (5.8%) | 9 (8.7%) | 9 (8.7%) | 0.050 |
| Valve intervention <sup>d</sup> | 11 (10.6%)<sup>a</sup> | 20 (19.2%)<sup>ab</sup> | 21 (20.2%)<sup>b</sup> | <0.001* |

\* Statistically significant overall change across the three time points ( $p < 0.05$ ) based on Cochran's Q test.
□, □: Time points with different superscript letters differed significantly in pairwise comparisons using McNemar's test, with a Bonferroni-corrected significance threshold of $p < 0.017$ .
<sup>c</sup>: Ablation was performed for atrial fibrillation, atrial flutter, ventricular tachycardia, or ventricular premature contractions.
<sup>d</sup>: Valve intervention includes MVR, AVR, TAVI, M-TEER, and other interventions for valvular disease.
Abbreviations: ABL, catheter ablation; AVR, aortic valve replacement; CABG, coronary artery bypass grafting; CRT-D, cardiac resynchronization therapy with defibrillator; ICD, implantable cardioverter-defibrillator; LVRR, left ventricular reverse remodeling; M-TEER, mitral transcatheter edge-to-edge repair; MVR, mitral valve replacement; PCI, percutaneous coronary intervention; TAVI, transcatheter aortic valve implantation.

### Factors associated with LVEF at Follow-up 2

In multivariable linear regression analysis, a higher ΔSimple-GDMT Score was independently associated with a higher LVEF at Follow-up 2 (β: 1.265, 95% CI: 0.237 to 2.293, p = 0.016). Baseline LVEF was positively associated with LVEF at Follow-up 2 (β: 0.349, 95% CI: 0.023 to 0.675, p = 0.036), whereas ischemic heart disease was negatively associated with LVEF at Follow-up 2 (β: −6.343, 95% CI: −11.487 to −1.198, p = 0.016). HF duration was also significantly associated with LVEF at Follow-up 2 (overall p = 0.001), with a significant nonlinear component (p = 0.014) **(Supplementary Table S2 in Online Resource 1)**.

### Clinical outcomes

Over a median follow-up of 33.8 months (mean 35.4 months), 52 patients (24.4%) died and 32 (15.0%) were hospitalized for HF. During follow-up, 108 patients were censored due to transfer of care to other hospitals. Kaplan–Meier analysis with pairwise log-rank tests using a Bonferroni-corrected significance threshold of p < 0.017 showed lower event rates for the primary composite outcome in both the Early LVRR group (p < 0.001) and the Late LVRR group (p = 0.009) compared with the No-LVRR group **(Figure 3a)**. For all-cause mortality, both the Early LVRR group (p < 0.001) and the Late LVRR group (p = 0.005) had fewer deaths than the No-LVRR group **(Figure 3b)**. For HF hospitalization, only the Early LVRR group had fewer events than the No-LVRR group (p = 0.002), whereas the Late LVRR group did not differ significantly (p = 0.132) **(Figure 3c)**.

**Figure 3:**
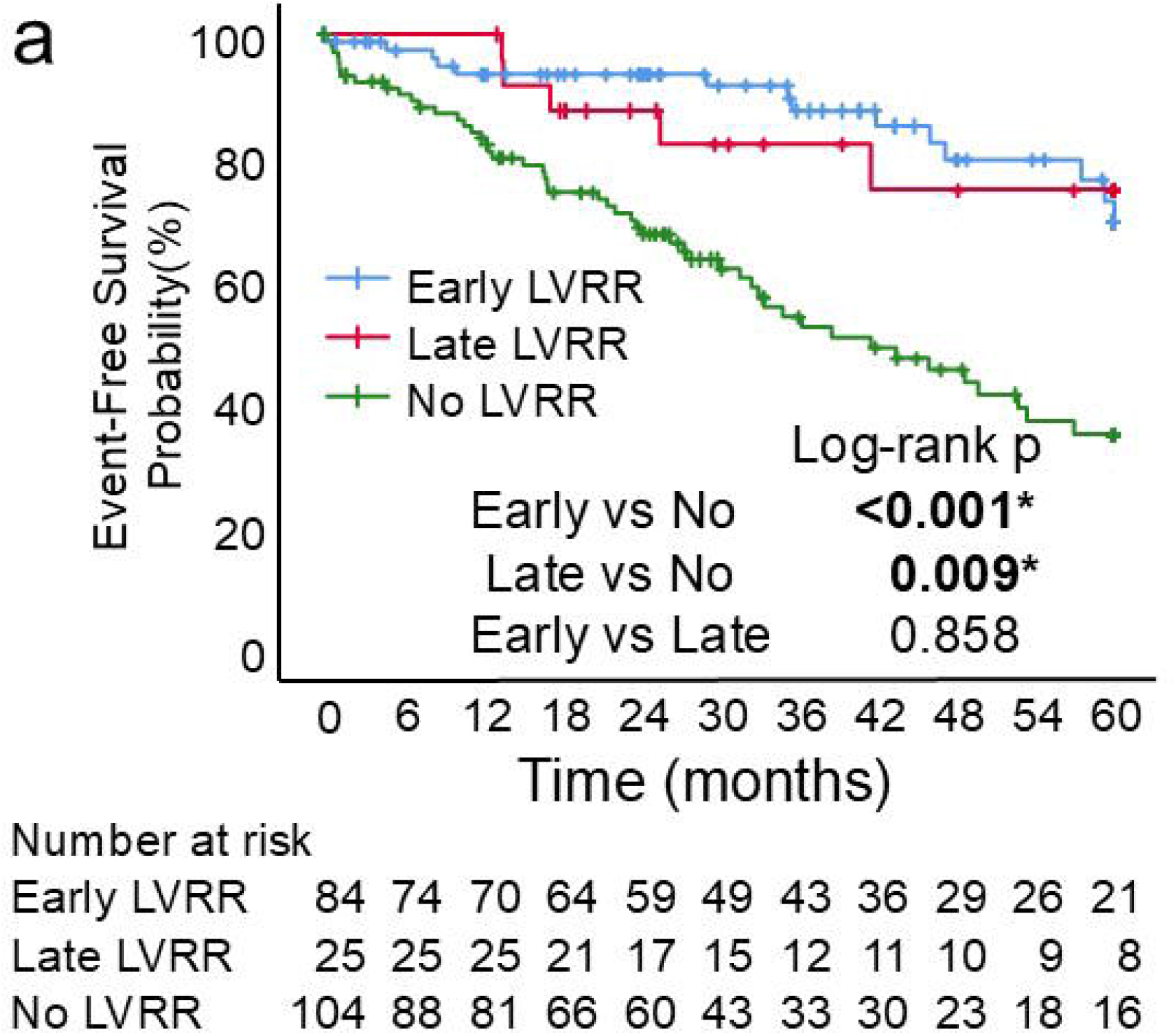
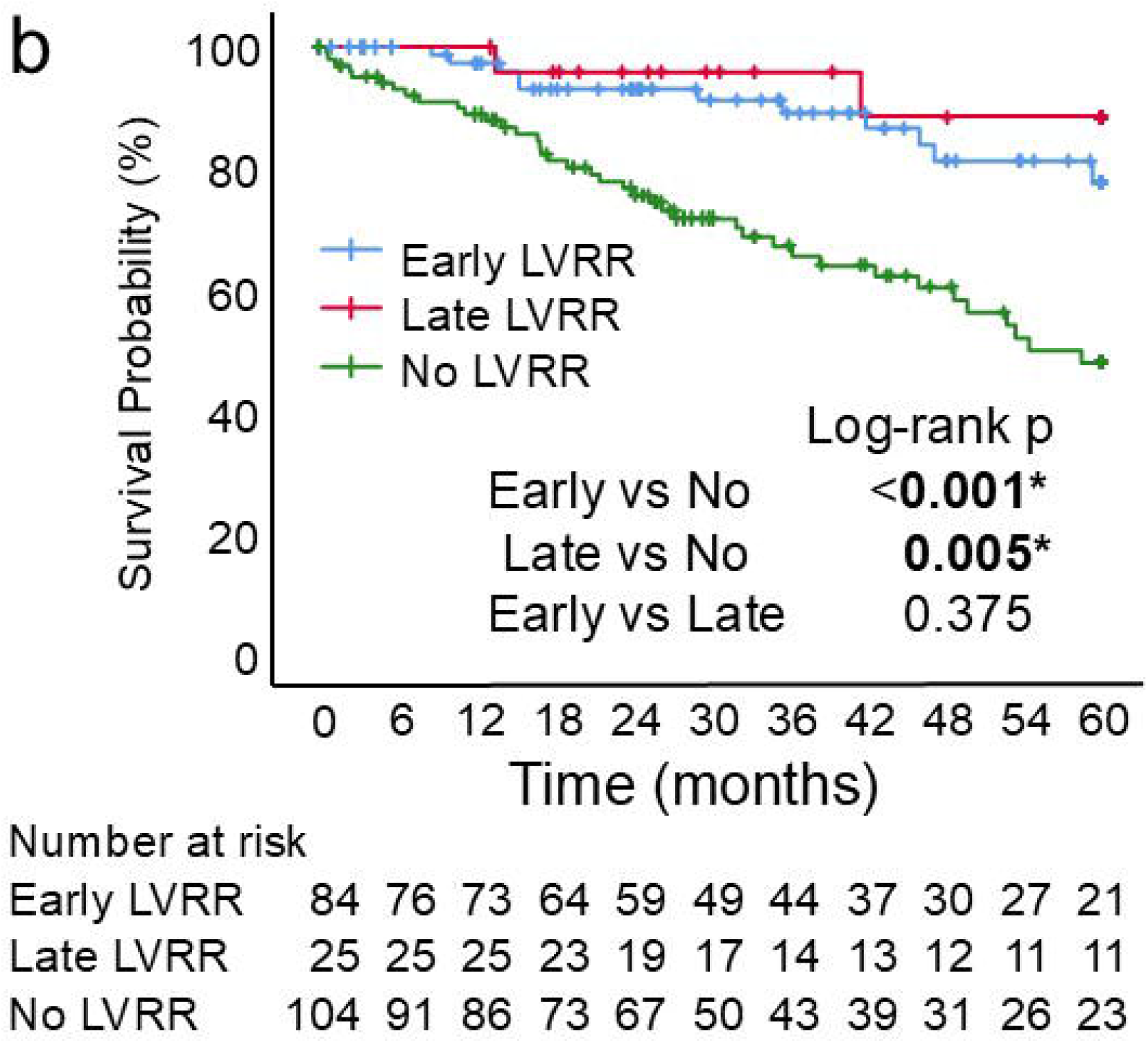
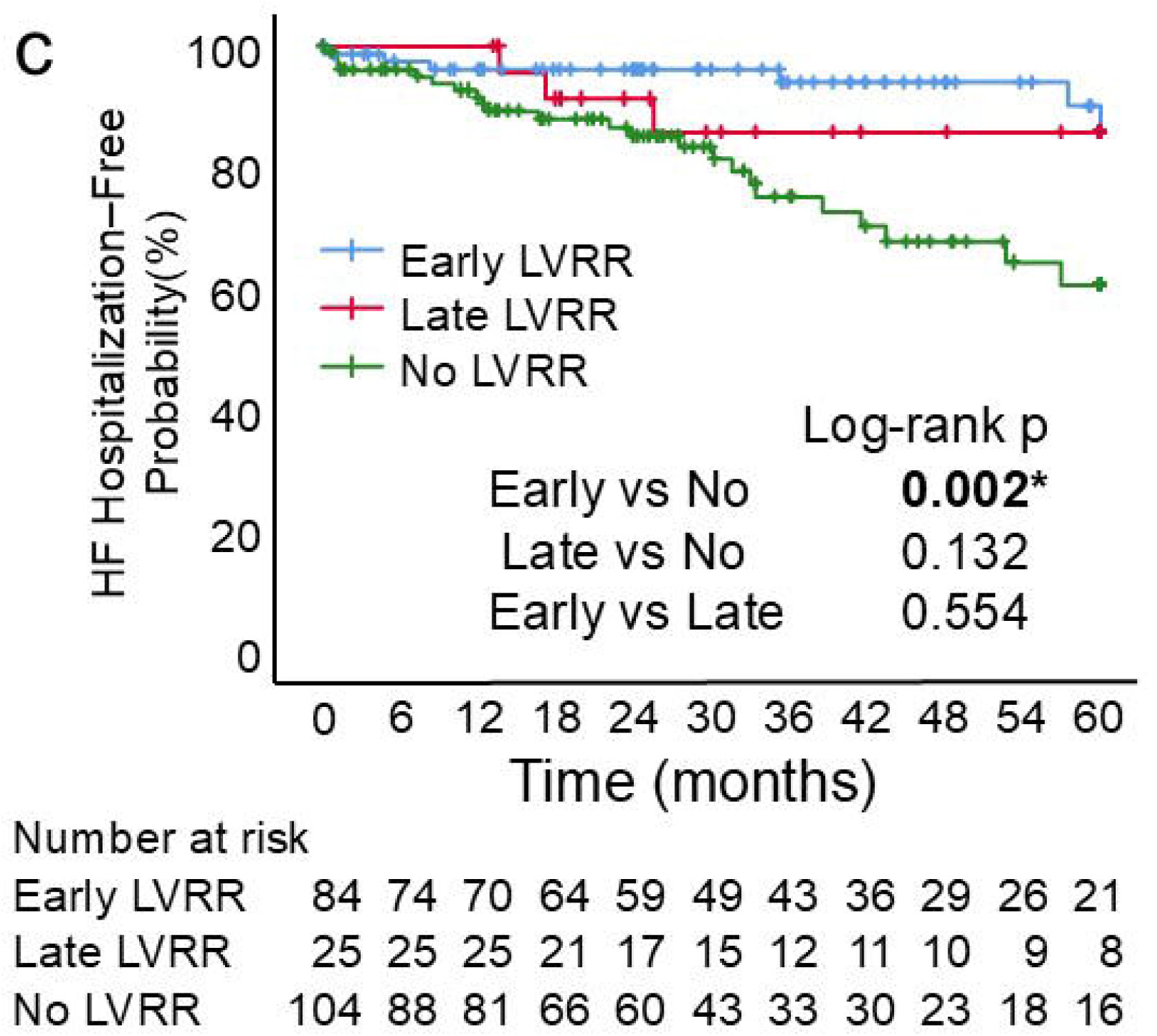
Kaplan–Meier curves for (a) the composite outcome of all-cause mortality and heart failure hospitalization, (b) all-cause mortality, and (c) heart failure hospitalization in the Early LVRR, Late LVRR, and No-LVRR groups Statistical significance for pairwise log-rank tests was determined using a Bonferroni-corrected threshold of p < 0.017. Abbreviations: HF, heart failure; LVRR, left ventricular reverse remodeling.

In multivariable Cox proportional hazards models **(Table 4)**, both Early and Late LVRR were consistently associated with a lower risk of the composite outcome compared with No-LVRR. In Model 1, the HRs were 0.299 (95% CI 0.164–0.544; p < 0.001) for Early LVRR and 0.293 (95% CI 0.115–0.744; p = 0.010) for Late LVRR. These associations remained consistent across Models 2–4, with HRs ranging from 0.263 to 0.307 for Early LVRR and from 0.343 to 0.370 for Late LVRR (all p < 0.05).

**Table 4.**
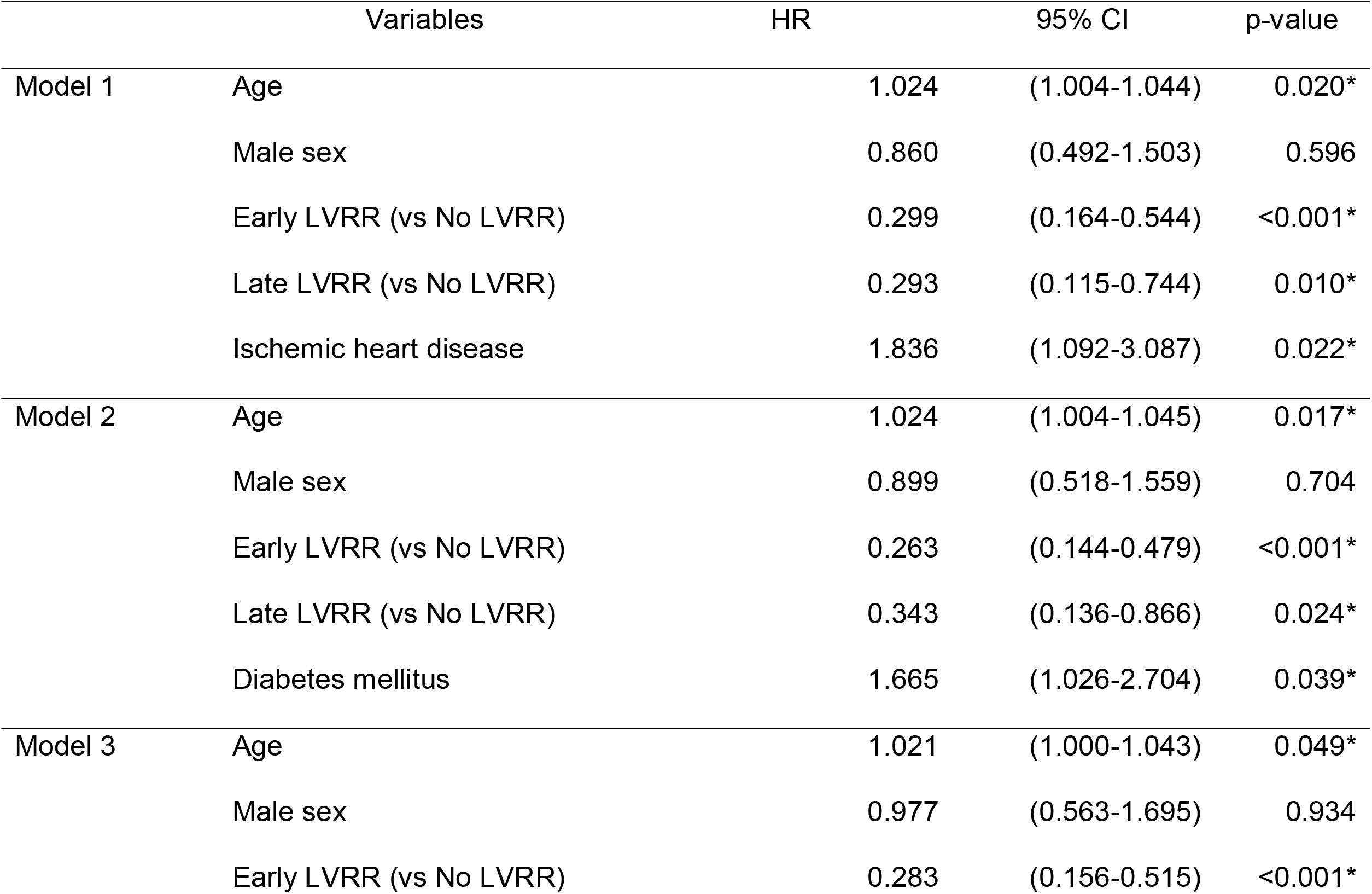
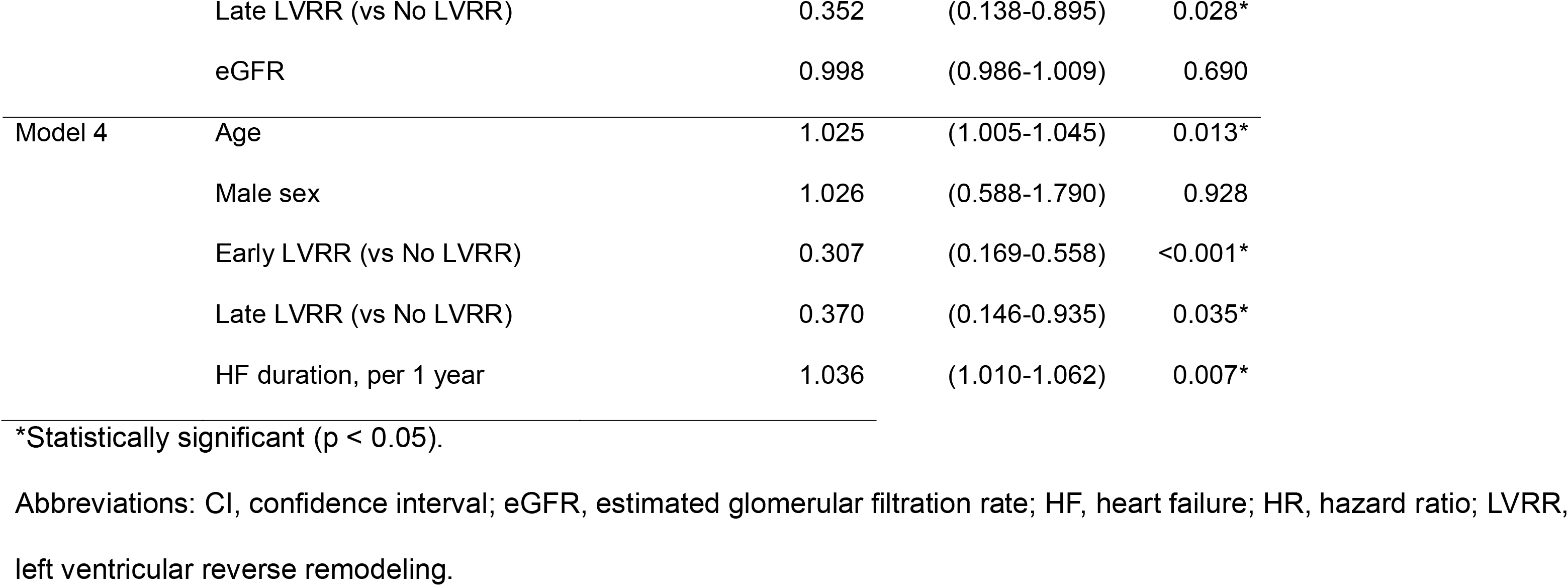
Multivariable Cox proportional hazards models for the composite outcome of all-cause mortality and HF.

|  | Variables | HR | 95% CI | p-value |
| --- | --- | --- | --- | --- |
| Model 1 | Age | 1.024 | (1.004-1.044) | 0.020* |
|  | Male sex | 0.860 | (0.492-1.503) | 0.596 |
|  | Early LVRR (vs No LVRR) | 0.299 | (0.164-0.544) | <0.001* |
|  | Late LVRR (vs No LVRR) | 0.293 | (0.115-0.744) | 0.010* |
|  | Ischemic heart disease | 1.836 | (1.092-3.087) | 0.022* |
| Model 2 | Age | 1.024 | (1.004-1.045) | 0.017* |
|  | Male sex | 0.899 | (0.518-1.559) | 0.704 |
|  | Early LVRR (vs No LVRR) | 0.263 | (0.144-0.479) | <0.001* |
|  | Late LVRR (vs No LVRR) | 0.343 | (0.136-0.866) | 0.024* |
|  | Diabetes mellitus | 1.665 | (1.026-2.704) | 0.039* |
| Model 3 | Age | 1.021 | (1.000-1.043) | 0.049* |
|  | Male sex | 0.977 | (0.563-1.695) | 0.934 |
|  | Early LVRR (vs No LVRR) | 0.283 | (0.156-0.515) | <0.001* |
|  | Late LVRR (vs No LVRR) | 0.352 | (0.138-0.895) | 0.028* |
|  | eGFR | 0.998 | (0.986-1.009) | 0.690 |
| Model 4 | Age | 1.025 | (1.005-1.045) | 0.013* |
|  | Male sex | 1.026 | (0.588-1.790) | 0.928 |
|  | Early LVRR (vs No LVRR) | 0.307 | (0.169-0.558) | <0.001* |
|  | Late LVRR (vs No LVRR) | 0.370 | (0.146-0.935) | 0.035* |
|  | HF duration, per 1 year | 1.036 | (1.010-1.062) | 0.007* |

\*Statistically significant ( $p < 0.05$ ).
Abbreviations: CI, confidence interval; eGFR, estimated glomerular filtration rate; HF, heart failure; HR, hazard ratio; LVRR, left ventricular reverse remodeling.

## Discussion

### Prognostic significance and clinical implications of late LVRR

LVRR is an established marker of favorable cardiac remodeling and is associated with a better prognosis in patients with HFrEF [20]. Although LVRR has most often been observed within the first 24 months in previous studies, our findings indicate that LVEF may continue to improve beyond this period. In our cohort, patients with Late LVRR showed progressive improvement in left ventricular function and dimensions after an initially modest response. Importantly, Late LVRR was associated with a lower risk of the composite outcome and all-cause mortality than No-LVRR. These findings extend previous observations in idiopathic dilated cardiomyopathy, in which late LVRR was associated with outcomes comparable to those of early LVRR [10], to a broader HFrEF population including ischemic and hypertensive etiologies. Although Late LVRR was associated with favorable overall outcomes, its association with heart failure hospitalization was less apparent. The mechanisms underlying differences in the timing of LVRR remain uncertain. Previous studies have demonstrated that LVEF trajectories vary over time and are associated with clinical outcomes [21, 22]. Differences in the underlying myocardial substrate and the longitudinal course of treatment may contribute to this heterogeneity. Further studies are needed to clarify whether the timing of LVRR varies according to HF etiology and underlying myocardial substrate, and whether the timing of LVRR provides additional information for long-term risk stratification.

### Clinical implications of longitudinal GDMT optimization

The longitudinal changes in GDMT may provide context for the different trajectories of LVRR observed in this study. Simple-GDMT Scores increased over time in all three groups, while the Late LVRR group showed continued intensification through Follow-up 2. In parallel, the patterns of non-pharmacological interventions differed across groups: several interventions increased during the early follow-up period in the Early LVRR group, whereas no significant increases in non-pharmacological interventions were observed between Follow-up 1 and Follow-up 2 in the Late LVRR group. In multivariable linear regression, greater increases in GDMT intensity were associated with higher LVEF at Follow-up 2. Because GDMT intensification and changes in LVEF were assessed over the same observation period, the direction of this association cannot be determined from our observational data. Nevertheless, the observed relationship suggests that the longitudinal pattern of GDMT intensification may be related to the timing and extent of functional improvement.

Implementation of GDMT in routine practice remains challenging because of multiple clinical and organizational barriers, including treatment complexity and concerns about hypotension, renal dysfunction, and hyperkalemia [23, 24]. Clinical inertia may also contribute to delayed or incomplete optimization [25]. This issue may be particularly relevant when HF care extends beyond specialized cardiology settings. In a survey using clinical vignettes, cardiologists were more likely than internal/family medicine physicians to modify GDMT [26]. Long-term HF management often extends across specialist and community care settings [27], and the concept of Late LVRR may provide a tangible clinical reference point that can be communicated across these settings. Recognition that meaningful improvement in left ventricular function may occur beyond the first 24 months may help maintain a shared focus on continued treatment optimization even when early improvement is limited. The potential value of this approach in supporting continued therapeutic engagement across different care settings warrants further investigation.

### Limitations

This study has several limitations. First, because of its observational design, causal relationships between GDMT intensification, non-pharmacological therapies, and LVRR cannot be established. Differences in treatment strategies and other unmeasured factors may also have influenced LVRR and clinical outcomes. Second, eligibility required echocardiographic follow-up beyond two years, which may have introduced selection bias by excluding patients with more advanced disease or early mortality. Third, baseline was defined as the first clinically available echocardiographic assessment at our institution rather than the onset or initial diagnosis of HF. Therefore, HF duration and treatment status at baseline may have varied among patients, and the terms “Early” and “Late” LVRR should be interpreted relative to this baseline TTE. Finally, although the Simple-GDMT Score captures prescribed drug classes and dose categories, it does not fully reflect actual treatment exposure, treatment duration, dose interruptions, or medication adherence.

## Conclusion

Late LVRR was associated with a lower risk of the composite outcome and all-cause mortality. Greater increases in GDMT intensity were associated with higher LVEF at follow-up, although causality cannot be established. Recognition of late functional improvement may provide a shared clinical reference point for long-term treatment optimization across diverse care settings.

## Data Availability

The data that support the findings of this study are not publicly available due to [ethical/privacy/legal] restrictions but are available from the corresponding author on reasonable request.

## Acknowledgments

This research did not receive any specific grant from funding agencies in the public, commercial, or not-for-profit sectors. During the preparation of this manuscript, the authors used ChatGPT (OpenAI) to improve grammar and language clarity. The authors reviewed and edited the content as needed and take full responsibility for the content of the published article.

