## Supplementary Material for "Prognostic impact of late left ventricular reverse remodeling in heart failure with reduced ejection fraction"

Heart and Vessels

Hayao Ikesugi, Shinya Fujiki, Satomi Kawai, Sho Hirayama, Soma Sato, Kazuyo Tanaka, Yuka Sekiya, Hiroki Tsuchiya, Takayuki Kumaki, Ryohei Sakai, Hiromi Kayamori, Tsugumi Takayama, Takeshi Kashimura, Takayuki Inomata

Department of Cardiovascular Medicine, Niigata University Graduate School of Medicine, Dentistry and Health Sciences

Corresponding Author: Shinya Fujiki

Online Resource 1. Supplementary tables and figures

**Supplementary Table S1. Echocardiographic parameters at Follow-up 1 and Follow-up 2 according to LVRR status**

|  |  | Overall (N = 213) | Early LVRR group | Late LVRR group | No-LVRR group | p-value |
| --- | --- | --- | --- | --- | --- | --- |
|  |  |  | (n = 84) | (n = 25) | (n = 104) |  |
| Follow-up 1 | |  |  |  |  |  |
|  | LVEF (%) | 38.1±13.7 | 51.9±7.6^a^ | 34.9±6.9^b^ | 27.6±7.7^c^ | <0.001* |
|  | LVDd (mm) | 56.0±10.2 | 49.7±6.6^a^ | 54.0±8.8^b^ | 61.5±9.8^c^ | <0.001* |
|  | LVDs (mm) | 45.7±11.2 | 36.5±5.9^a^ | 44.8±7.9^b^ | 53.4±9.4^c^ | <0.001* |
| Follow-up 2 | |  |  |  |  |  |
|  | LVEF (%) | 40.0±15.2 | 52.7±10.6^a^ | 49.1±6.4^b^ | 27.5±8.2^c^ | <0.001* |
|  | LVDd (mm) | 55.5±10.2 | 49.6±6.5^a^ | 49.6±7.4^a^ | 61.7±9.6^b^ | <0.001* |
|  | LVDs (mm) | 45.0±12.1 | 36.1±7.1^a^ | 38.1±6.6^a^ | 53.8±9.7^b^ | <0.001* |

* Statistically significant difference (p < 0.05) based on Kruskal–Wallis test.

ᵃ, ᵇ, ^c^: Groups with different superscript letters differed significantly in pairwise comparisons using the Mann–Whitney U test, with a Bonferroni-corrected significance threshold of p < 0.017.

Abbreviations: LVDd, left ventricular end-diastolic diameter; LVDs, left ventricular end-systolic diameter; LVEF, left ventricular ejection fraction; LVRR, left ventricular reverse remodeling.

**Supplementary Table S2. Multivariable linear regression analysis for LVEF at Follow-up 2**

|  | Model 1 β (95% CI) | p-value | Model 2 β (95% CI) | p-value |
| --- | --- | --- | --- | --- |
| ΔSimple-GDMT Score ^a^, per 1 point | 1.471 (0.418 to 2.524) | 0.006* | 1.265 (0.237 to 2.293) | 0.016* |
| Baseline LVEF, per 1% | 0.267 (−0.046 to 0.581) | 0.094 | 0.349 (0.023 to 0.675) | 0.036* |
| Age, per 1 year | −0.102 (−0.270 to 0.067) | 0.236 | −0.074 (−0.247 to 0.100) | 0.403 |
| Male sex | −3.688 (−8.949 to 1.573) | 0.168 | −2.089 (−7.267 to 3.089) | 0.427 |
| Ischemic heart disease | — | — | −6.343 (−11.487 to −1.198) | 0.016* |
| Log-transformed BNP | — | — | −1.511 (−3.600 to 0.577) | 0.155 |
| eGFR, per 1 mL/min/1.73 m² | — | — | −0.003 (−0.087 to 0.082) | 0.951 |
| HF duration ^b^, overall | — | — | — | 0.001* |
| Nonlinear component | — | — | — | 0.014* |

Model 1 was adjusted for baseline LVEF, age, and sex. Model 2 was additionally adjusted for ischemic heart disease, log-transformed BNP, eGFR, and HF duration. β coefficients are presented with 95% confidence intervals.

* Statistically significant (p < 0.05).

^a^: Simple-GDMT Score is a composite score reflecting the use and dose of four classes of GDMT (β-blocker, ACEi/ARB/ARNi, MRA, and SGLT2i; score range, 0–9). ΔSimple-GDMT Score indicates the change from baseline to Follow-up 2.

^b^: HF duration was modeled using a restricted cubic spline with three knots; therefore, a single β coefficient and 95% confidence interval are not presented.

Abbreviations: ACEi, angiotensin-converting enzyme inhibitor; ARB, angiotensin II receptor blocker; ARNi, angiotensin receptor–neprilysin inhibitor; BNP, B-type natriuretic peptide; eGFR, estimated glomerular filtration rate; GDMT, guideline-directed medical therapy; IHD, ischemic heart disease; LVEF, left ventricular ejection fraction; MRA, mineralocorticoid receptor antagonist; SGLT2i, sodium–glucose cotransporter 2 inhibitor.

**Supplementary Figure S1. Temporal trends in the implementation rates of each drug class: (A) β-blocker, (B) ACEi/ARB, (C) ARNi, (D) MRA, and (E) SGLT2i**

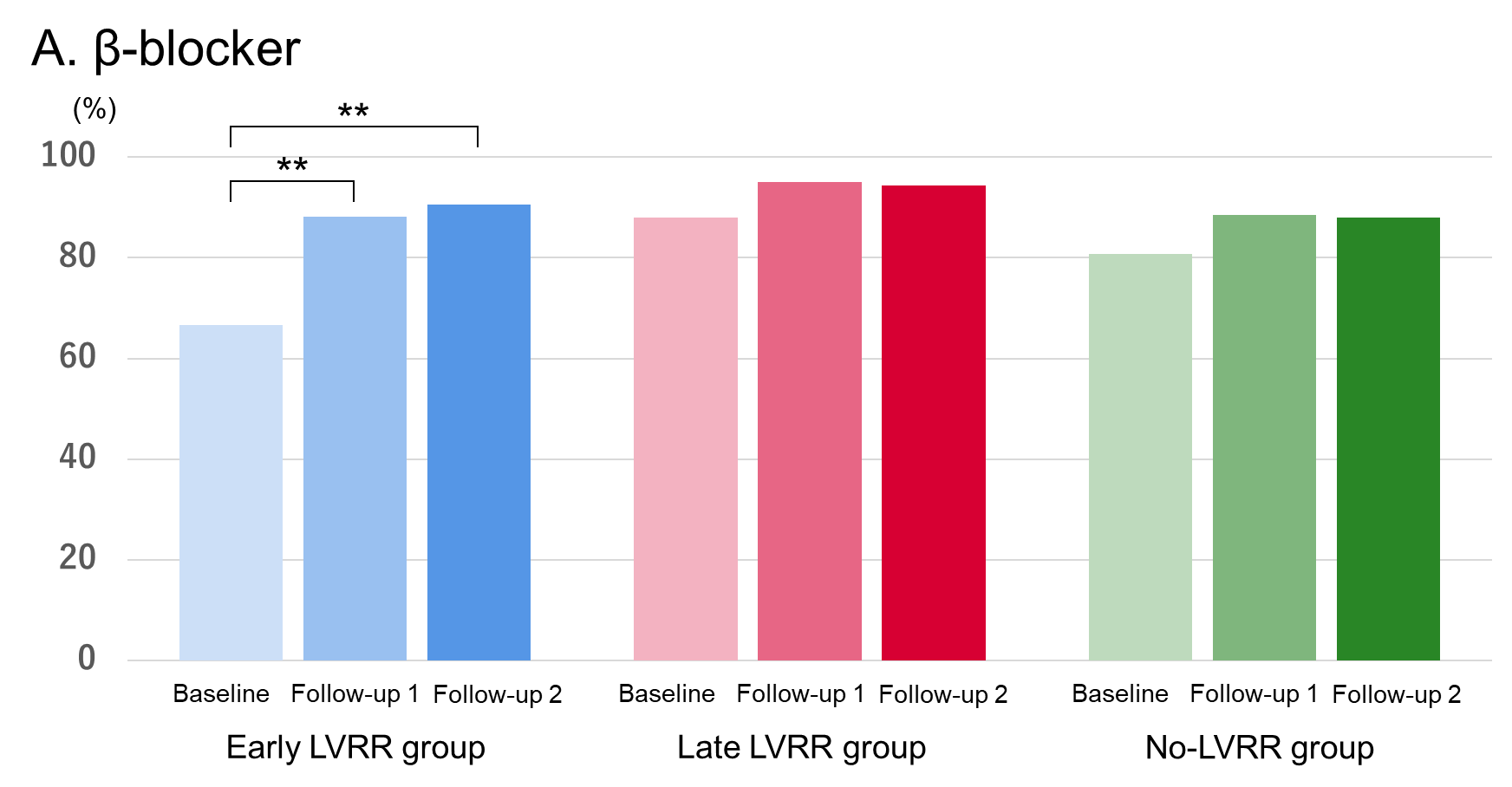

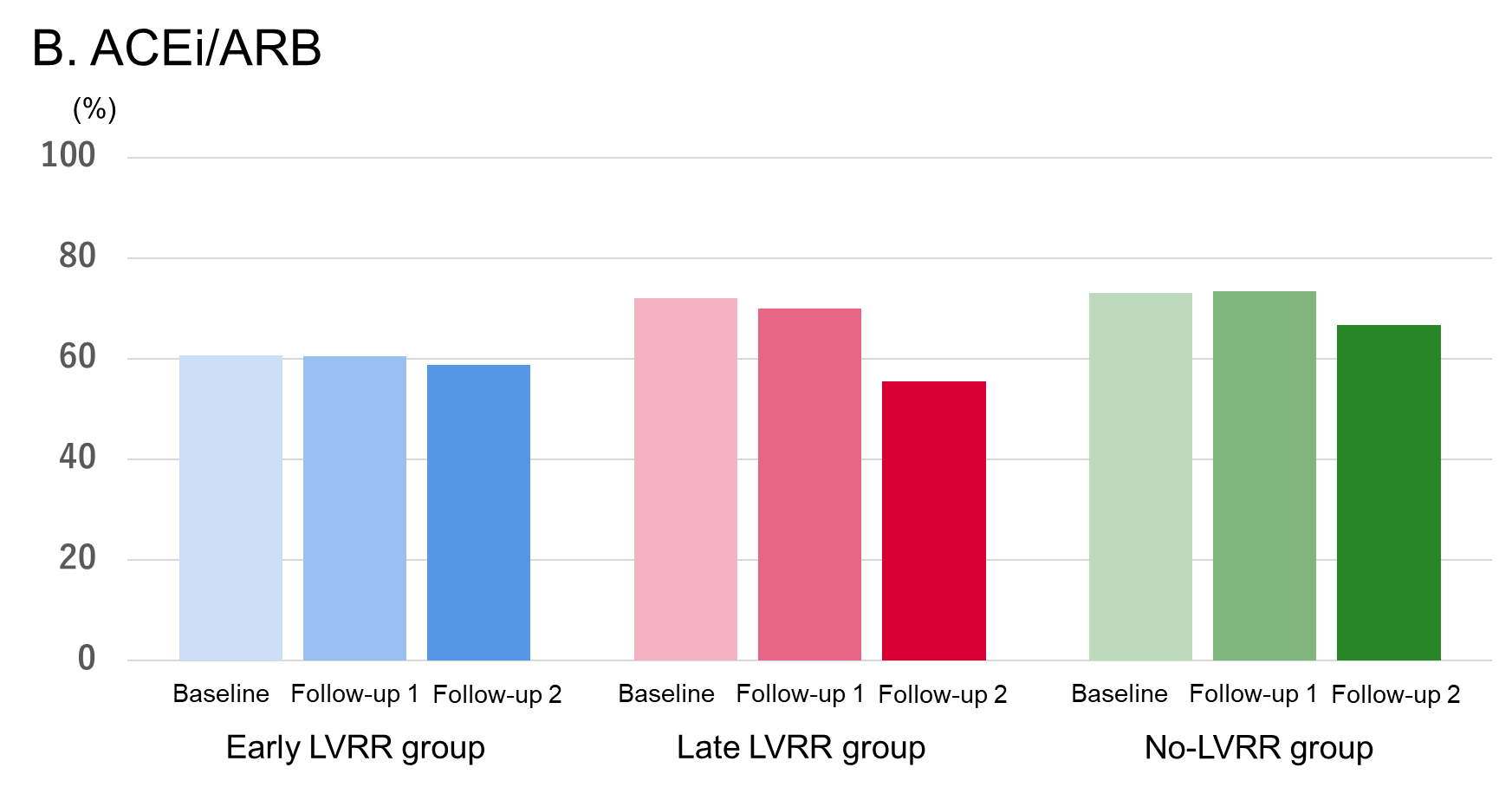

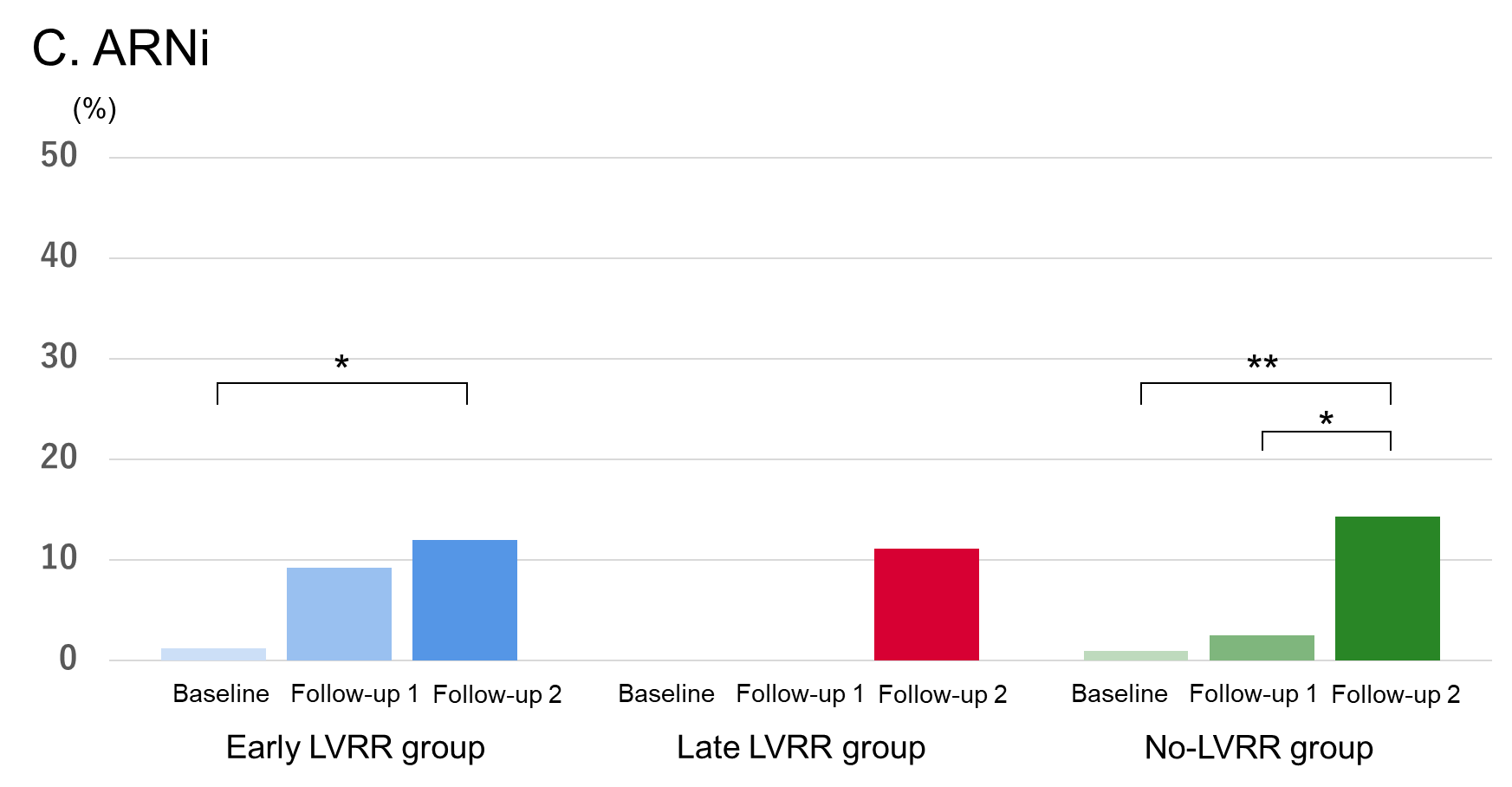

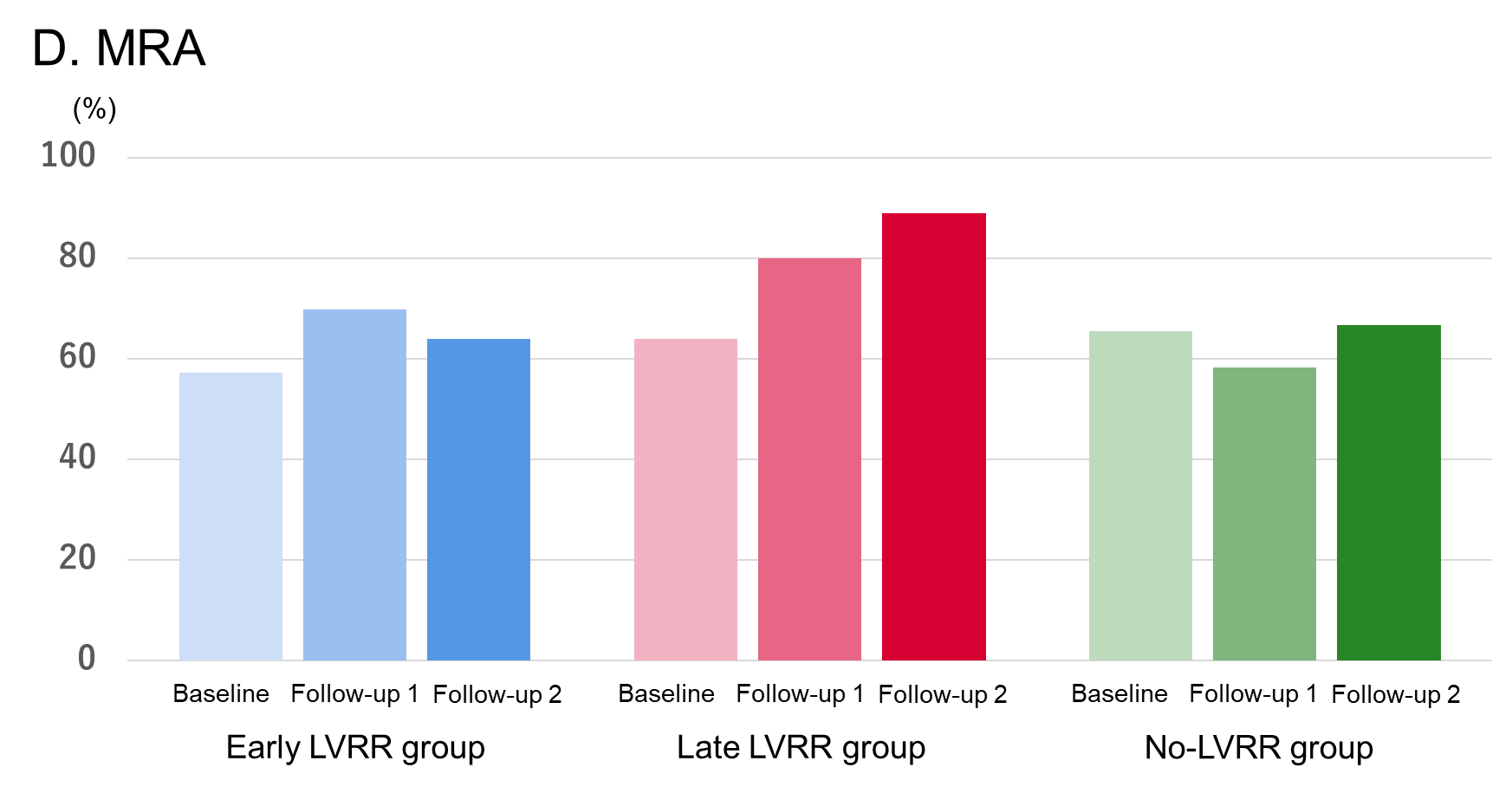

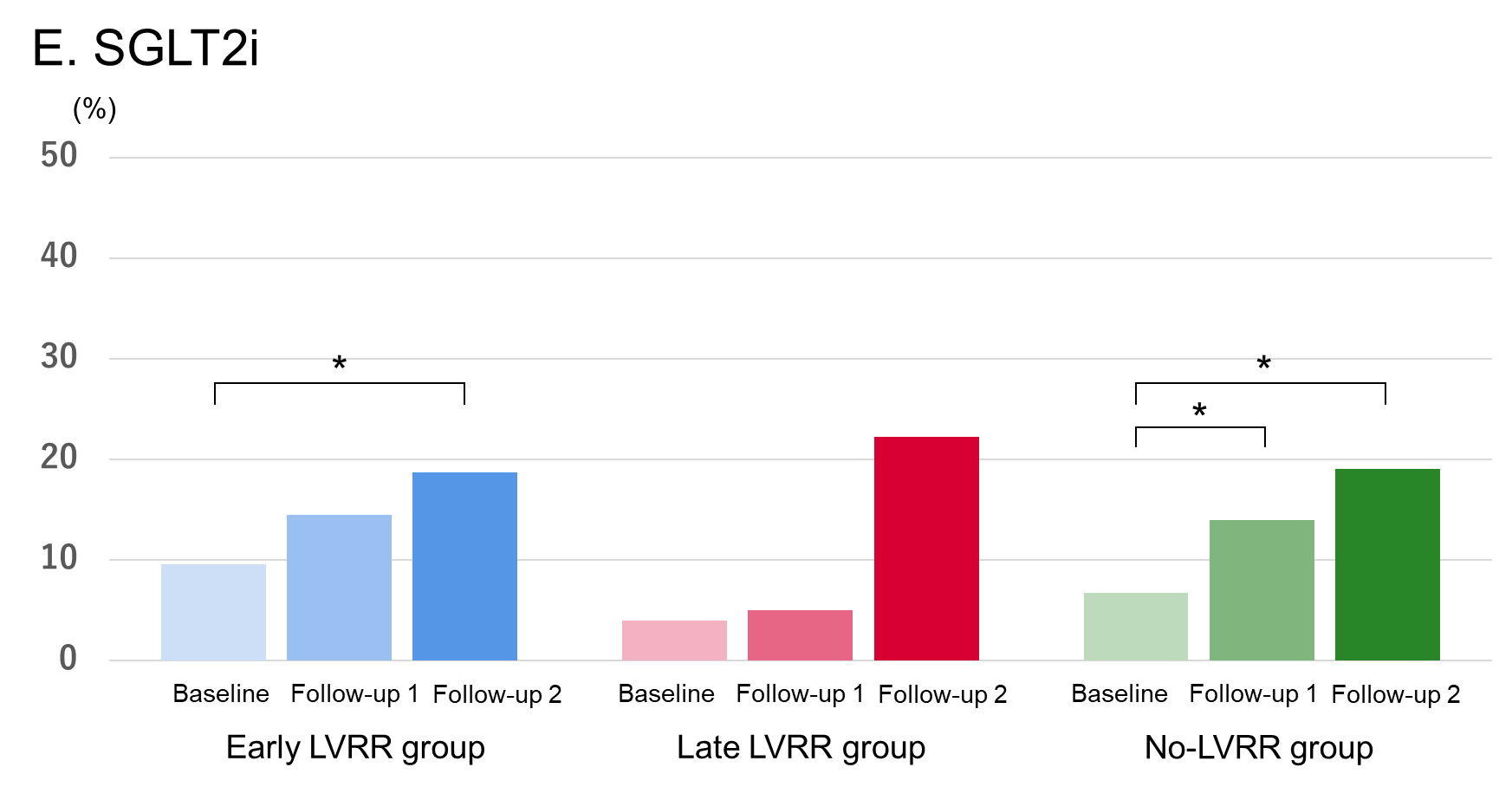

Blue bars represent the Early LVRR group, red bars the Late LVRR group, and green bars the No-LVRR group. Each bar indicates the implementation rates of each drug class at baseline, Follow-up 1 (6 to <24 months), and Follow-up 2 (24–78 months).

*p < 0.017, **p < 0.0033. Statistical significance was determined using Bonferroni-corrected thresholds for three pairwise comparisons.

Abbreviations: ACEi, angiotensin-converting enzyme inhibitor; ARB, angiotensin II receptor blocker; ARNi, angiotensin receptor–neprilysin inhibitor; LVRR, left ventricular reverse remodeling; MRA, mineralocorticoid receptor antagonist; SGLT2i, sodium–glucose cotransporter 2 inhibitor.

**Supplementary Figure S2. Temporal changes in (A) loop diuretic prescription rates and (B) furosemide-equivalent doses**

**A. loop diuretic prescription rates**

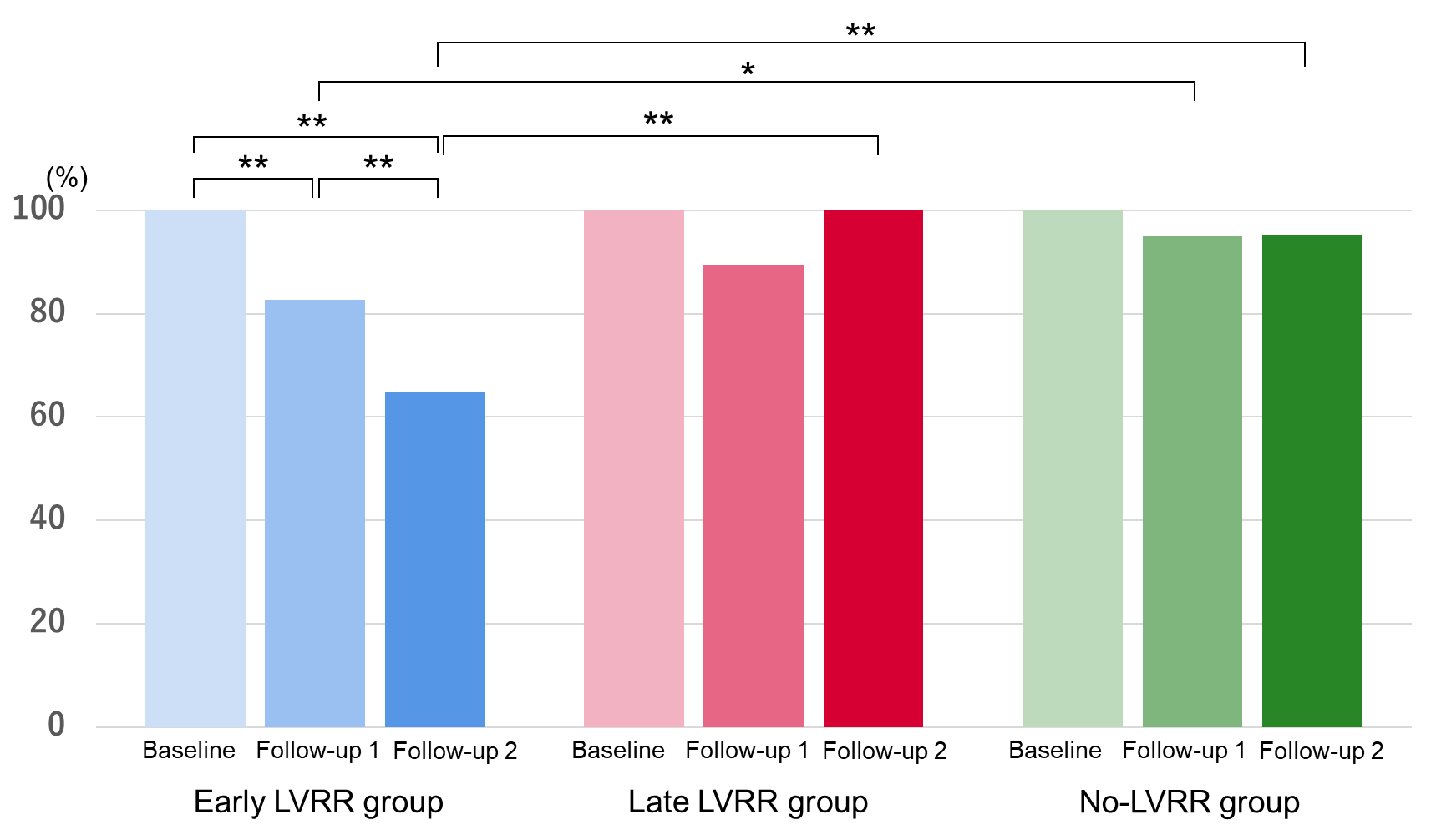

**B. furosemide-equivalent doses**

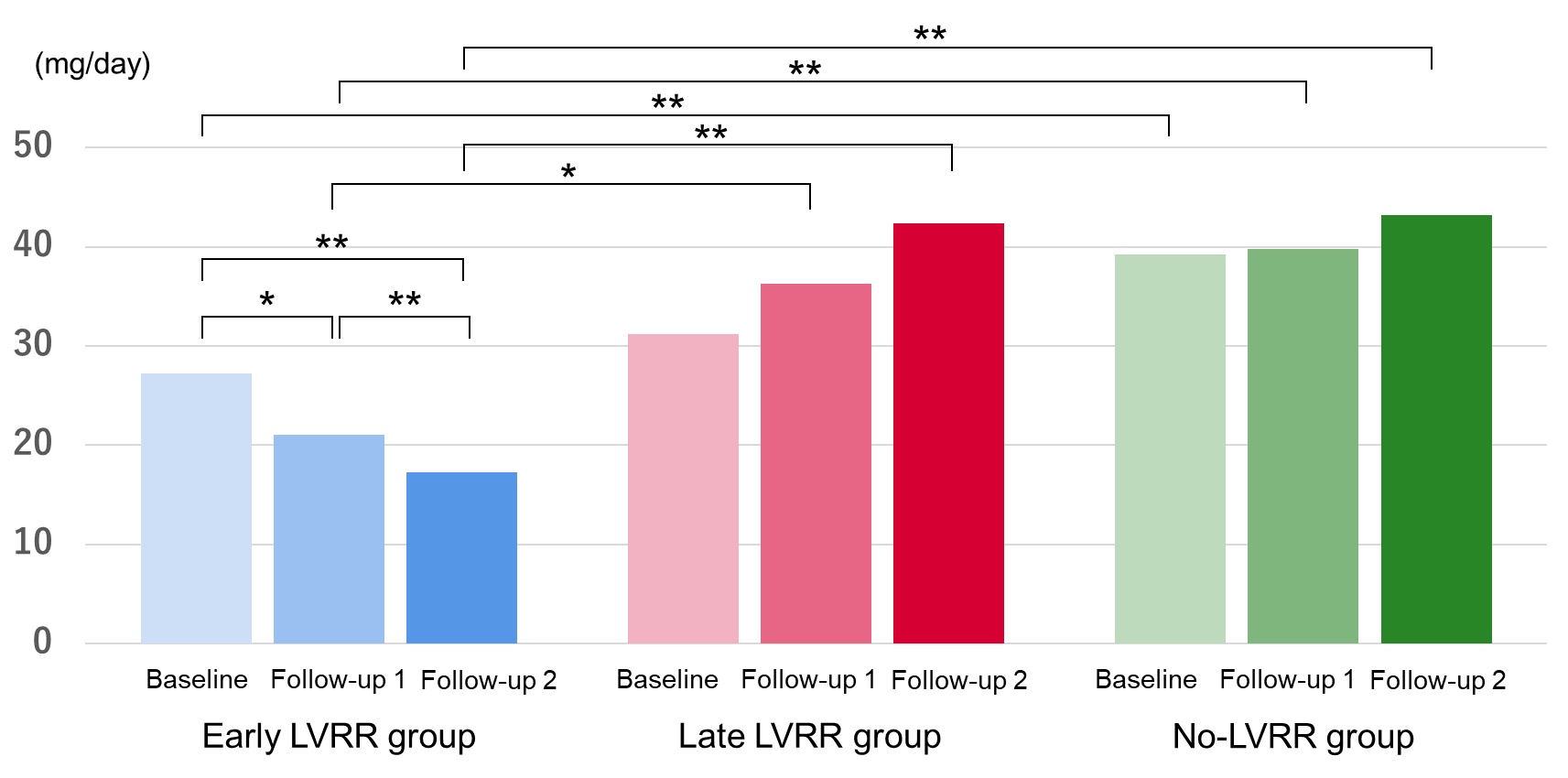

Blue bars represent the Early LVRR group, red bars the Late LVRR group, and green bars the No-LVRR group. The bars show loop diuretic prescription rates and furosemide-equivalent doses at baseline, Follow-up 1 (6 to <24 months), and Follow-up 2 (24–78 months).

*p < 0.017, **p < 0.0033. Statistical significance was determined using Bonferroni-corrected thresholds for three pairwise comparisons.
